# Shared predicted effector genes of insomnia, anxiety and depression implicate synaptic processes as transdiagnostic drug targets

**DOI:** 10.1101/2025.10.18.25338281

**Authors:** Marijn Schipper, Alexey A. Shadrin, Cato Romero, Eleni Friligkou, Renato Polimanti, Danielle Posthuma, Eus J.W. Van Someren, Elleke Tissink

## Abstract

**Background:** Insomnia (INS) is a major factor determining the risk, severity, treatment resistance and relapse of depression (DEP) and anxiety disorders (ANX), suggesting shared neurobiological vulnerability. Identifying common biological mechanisms that are therapeutically actionable is a foundational step towards improving current clinical care. Since INS, DEP, and ANX share substantial genetic risk, pinpointing the specific genetic factors that influence these three conditions simultaneously offers a route to discover pharmacological targets with transdiagnostic potential.

**Methods:** We propose a framework of multi-level trivariate genetic analysis to identify the shared genetic component of INS, DEP and ANX using genome-wide association studies (GWAS). We trace shared components through trivariate genetic overlap, local genetic correlations, overlapping genome-wide significant loci, shared predicted effector genes, and shared putative causal variants.

**Results:** We show that INS, DEP and ANX share 55% of their GWAS signal. We identify 195 cross-disorder loci, many of which are likely arising from the same causal variant (at least 50%) or predicted effector gene (60-80%). We find convergence of shared predicted effector genes on biological processes such as inhibitory synaptic transmission, neuronal organization, and axonal development. The majority of shared predicted effector genes that are also druggable are involved in synaptic transmission and located in synaptic membranes, nominating *CACNA2D3, DRD2, GRIA1* and *GRM5* as key targets for drug development to treat comorbid INS, DEP and ANX.

**Conclusion:** These results reveal an interconnected but mechanistically diverse basis for shared genetic liability across co-occurring INS, DEP and ANX, and offer potential candidates for future pathway-tailored transdiagnostic therapeutic targets.

## Introduction

One-third of the general population has difficulties falling or staying asleep, which can severely impact quality of life when manifesting frequently and persistently as insomnia disorder (INS)^1^. INS may develop as a symptom of conditions such as depression (DEP) or anxiety disorders (ANX), or can alternatively precede and increase the risk of developing these conditions^2,3^. Besides high rates of lifetime multimorbidity^4^, INS, DEP and ANX show transdiagnostic intervention efficacy^5^ and share multimodal neuroimaging correlates^6^. Quantitative modelling in family and twin studies has consistently found that comorbidity between DEP and ANX^7^ and INS and DEP^8^ is more plausibly explained by shared risk factors or bidirectional causality than symptom overlap or diagnostic imprecision. Identifying the mechanism(s) of common neurobiological vulnerability may guide the development of transdiagnostic treatments, potentially improving outcomes across multiple conditions simultaneously^9,10^.

Novel pharmacological developments are about 2.6 times more successful when supported by genetic evidence^11^, yet the molecular genetic risk factors specifically shared across the triad of INS, DEP, and ANX have not been studied systematically to date. Based on decades of twin research^12^, it is widely recognized that the heritability of INS (39%^13^), DEP (37%^14^), ANX (32%^15^) is largely shared (>50% for DEP, complete for ANX). More recently, the molecular genetic signals of INS^16^, ANX^17,18^, and DEP^19,20^ have been intensively investigated in genome-wide association studies (GWAS). These studies identified common genetic variants (i.e. single nucleotide polymorphisms; SNPs) that are significantly over-or underrepresented in patients living with one of these conditions. The phenotypic variance in INS, ANX, or DEP that can be explained by additive effects of such common variants (i.e. observed SNP-heritability, 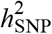) is modest (6-7%^16,17,20^), and is estimated to involve ~12,000 putatively causal SNPs with fairly low effect sizes compared to other psychiatric disorders^21^.

The degree to which these SNPs overlap and their effect sizes correlate across conditions (i.e. genetic overlap and genetic correlation [*r*_g_] respectively) can be a statistical byproduct of methodological artefacts (i.e. phenotype misclassification^22^) or point to real shared genetic risk. Previous pairwise studies show extensive genetic correlation and overlap between DEP-ANX (*r*_g_=.87^23^, and 85-95% of putative causal SNPs shared, with 82-91% sign concordance^17,20,24^). Comparisons with INS are often omitted^23,25,26^, but two bivariate studies report the genetic overlap between DEP-INS to be 65%^27,28^, and one cross-disorder study estimates the pairwise genetic correlations of INS-DEP and INS-ANX to be *r*_g_=.35 and .45 respectively^21^. These non-zero genetic correlations indicate the presence of pleiotropy^29–31^, i.e. the statistical association of one SNP or gene with multiple traits, but do not pinpoint where these SNPs or genes are located or what their (potentially targetable) mechanism of action is.

Identifying specific genomic regions that impact cross-disorder liability has been challenging in the past due to insufficient statistical power^21^. Fortunately, with sample sizes growing exponentially in psychiatric genetic studies^32^, we can now leverage recently published GWAS of INS^16^, ANX^17^, and DEP^19,20^ to conduct well-powered local genetic correlation analyses. Importantly, the odds of successful drug development improve with increasing genetic evidence that the target gene is causal^11^. Colocalization analysis^33^ and effector gene prediction^34^ can be used to filter cross-disorder loci for those that are driven by a shared putative-causal variant or gene. This enables to subsequently trace down onto which biological mechanisms and drug targets with transdiagnostic effect likely-causal genetic effects converge.

We propose a multi-level analysis framework, combining existing methods, to discover shared biological pathways with genetically-supported targets for potential transdiagnostic treatment across INS, DEP and ANX. At the genome-wide level, we estimate the trivariate genetic overlap across conditions to be 55%. We subsequently identify 195 unique loci with shared genome-wide significance or correlating genetic associations for at least two traits. We then show that at least 50% and 60-80% of these loci also share the same putative-causal variant or predicted effector gene respectively. These shared predicted effector genes are almost exclusively involved in inhibitory synaptic transmission and organization, and neuronal and axonal development. We find that the majority of our shared predicted effector genes that have previously been investigated or approved as drug targets are in fact indicated for INS, DEP and/or ANX, many of which are involved in the same synaptic transmission pathways. Altogether, this study shows that many genomic regions associated across INS, DEP and ANX map to the same putative-causal variants and genes, that converge on shared biological pathways being potentially therapeutically actionable.

## Methods and Materials

### GWAS summary statistics

We obtained access to GWAS summary statistics of INS^16^, ANX^17^, and DEP^19,20^, originally including participants from UK Biobank, 23andMe Research Institute, Million Veteran Program, Psychiatric Genetics Consortium, iPSYCH, FinnGen, and All of Us. Cross-ancestry summary statistics were available for ANX and DEP, including individuals from European (EUR), South Asian (SAS), East Asian (EAS), and African (AFR) genetic ancestries. Only EUR individuals were included in the original GWAS of INS and compared to the EUR subset of ANX and DEP where necessary. To examine the impact of broad phenotype definitions and sample overlap due to the inclusion of UKB across GWAS summary statistics, a subset of the EUR summary statistics without UK Biobank were used for sensitivity analyses (Supplementary Note 1).

In accordance with recent methodological recommendations^35^, effective sample size (*N*_eff_) of each trait was first calculated for each individual sample included in the GWAS meta-analysis and subsequently summed. We defined genome-wide significant loci according to the FUMA definition^36^.

### Genome-wide genetic overlap

We applied Trivariate MiXeR^37^ to disentangle the pattern of polygenic overlap among the EUR GWAS summary statistics. This recently released tool (https://github.com/precimed/mix3r) allows for sample overlap and uses 489 unrelated subjects of European ancestry from 1000 Genomes phase 3 as LD reference panel^38^ including SNPs with a minor allele frequency (MAF)>.01 (*n*_SNPs_=9,997,231). Following recommendations from the authors, we ignored all inter-variant correlations with *r*^2^<0.05 (considering them to be very noisy in a sample of 489 subjects) and excluded the MHC region (CHR6:BP25000000-34000000, GRCh37/hg19 build)^39^. The analysis was restricted to overlapping SNPs across summary statistics (N_SNPs_=3,122,674). Output parameters such as the proportion of non-null SNPs (polygenicity; *π*) where multiplied by the total number of SNPs in the LD reference panel for interpretability.

### Local genetic correlations

LAVA (Local Analysis of [co]Variant Association)^41^ was used for local *r*_g_ analysis, i.e. detecting genomic regions of shared association between our three phenotypes. Sample overlap was estimated with LDSC and the analysis was restricted to overlapping SNPs across summary statistics. We used 2,495 predefined loci that are roughly independent and the 1000 Genomes reference data for analysis with EUR GWAS summary statistics as provided (https://github.com/josefin-werme/LAVA). Using the AFR GWAS was not feasible due to the absence of suitable reference panels. SAS and EAS summary statistics did not have sufficient statistical power to run this analysis. For each region, traits devoid of local heritability (*p*<1×10^−4^, a threshold used in previous work^42–44^) were filtered out before estimating bivariate *r*_g_. This resulted in 580 EUR bivariate tests performed. We corrected the alpha (α) level for statistical significance of local bivariate *r*_g_ to α=(.5/582)=8.59×10^−5^.

### Shared putative causal SNPs

We ran COLOC (v5.2.3^45^) under a single variant assumption, in all overlapping LAVA locus pairs and GWS locus pairs. When only Z-scores were available, effect sizes (*β*) and standard errors (*SE*) were estimated from Z-scores (*Z*), effective sample size (*N)* and the minor allele frequency (*p*) as:

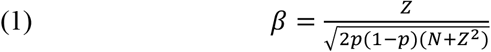

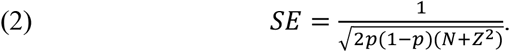

We deemed every locus-pair with a posterior probability >.5 for the case that both traits are associated and share a single causal variant to be significantly colocalizing (H4). As a sensitivity analysis we used the SuSiE COLOC implementation that allows for multiple shared variants^46^, with the 1000Genomes European ancestry individuals as LD reference^47^.

### Shared predicted effector genes

We applied FLAMES (Fine-mapped Locus Assessment Model of Effector geneS)^34^ to identify the most likely-causal gene in each shared locus identified in our study. Throughout this manuscript “predicted effector gene” is used to refer to the genes that mediate the observed variant-level associations observed in GWAS. FLAMES (Supplementary Note 2) was applied to each condition (INS, ANX, DEP) with the available EUR and cross-ancestry summary statistics (https://github.com/Marijn-Schipper/FLAMES). In short, FLAMES integrates SNP-to-gene evidence and convergence-based evidence into a single prediction for each fine-mapped GWAS signal. Note that loci can be left unresolved by FLAMES in the case of (1) unsuccessful fine-mapping due to complex LD (e.g. MHC region), (2) no proximal genes (<750 kilobases) from the credible set, or (3) an estimated cumulative precision <0.75.

### Pathway analysis

We performed pathway analyses to find patterns of enrichment of effector genes shared across INS, DEP and ANX, and of effector genes uniquely shared across two traits. As previously described in the FLAMES paper^34^, pathway membership of genes was encoded in a binary vector, and FLAMES prioritized genes were also encoded in a binary vector. We subsequently performed logistic regression, regressing the FLAMES genes of a phenotype on the pathways. Pathways were defined as Gene Ontology (GO)^48^ Biological Processes and Cellular Components (*n*=8,760) from MSigDB (v.2023.1). We corrected for multiple testing (α=(.05/8,760)/3=1.90×10^−6^) and required an overlap of ≥2 genes.

### Finding drug targets for predicted effector genes and mapping them to indications

To identify currently druggable shared effector genes, we downloaded the OpenTargets known drug dataset (version 25.12; https://ftp.ebi.ac.uk/pub/databases/opentargets/platform/25.12/output/known_drug/). We extracted all genes with an indication for anxiety, insomnia or depression. We manually cross-referenced the 43 drug-target pairs in the DrugBank database^49^, noting whether the drug had been approved or investigated in clinical trials for any of our phenotypes of interest.

## Results

Our analyses are based on previously published GWAS summary statistics for ANX^17^, DEP^19,20^ and INS^16^, (Supplementary Table 1, Supplementary Note 1). The original studies had an effective sample size of *N*_INS_=1,114,549, *N*_ANX_=529,764, *N*_DEP_=1,061,269 based on individuals of European (EUR) and African (AFR; *N*_ANX_=80,104; *N*_DEP_=132,002) genetic ancestry, and also performed cross-ancestry meta-analyses (*N*_ANX_=627,041; *N*_DEP_=1,118,664). With these data, we investigated the genetic relationship between INS, ANX and DEP on a genome-wide, locus, and causal gene/variant level.

**Table 1.**
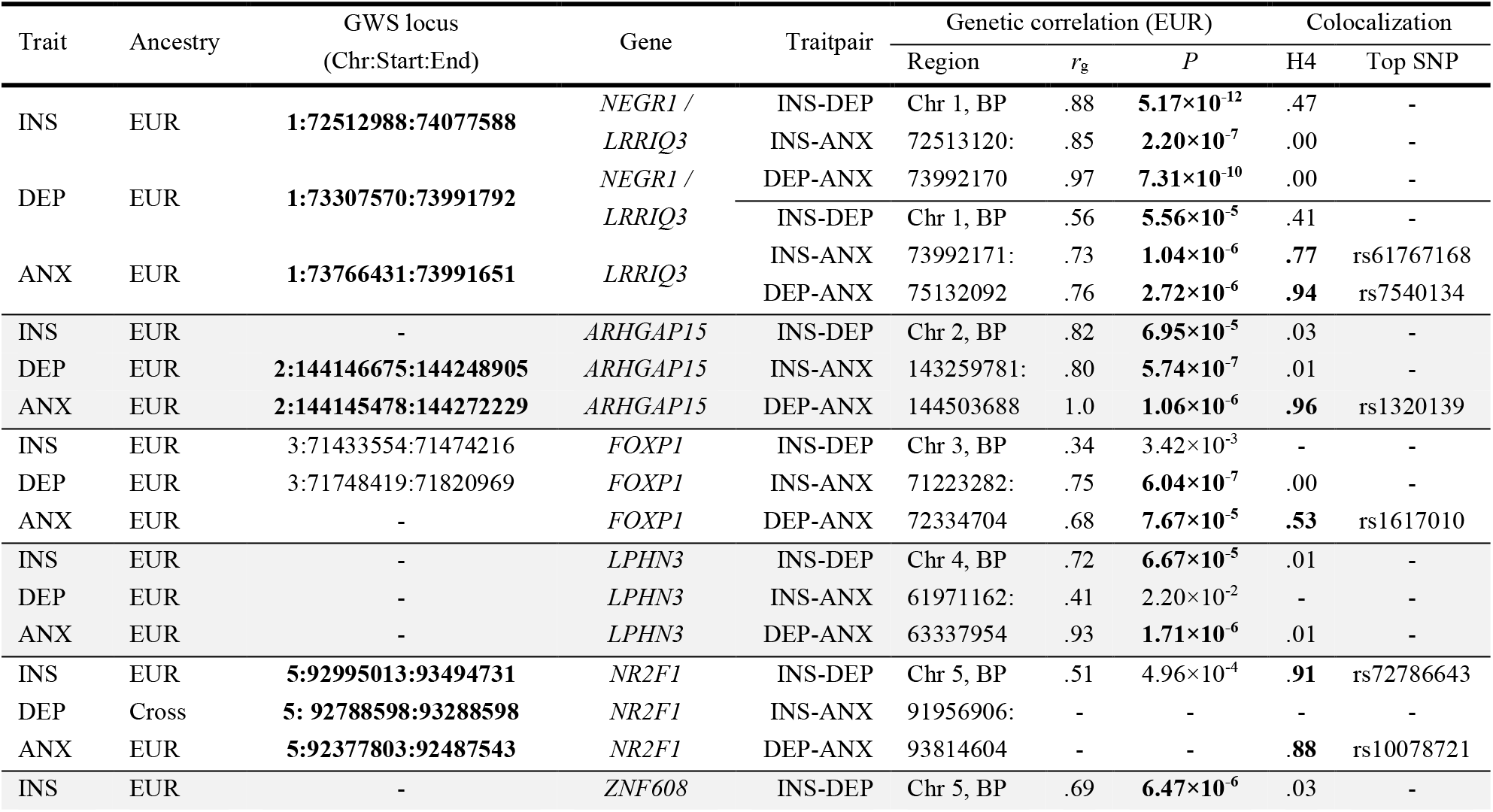
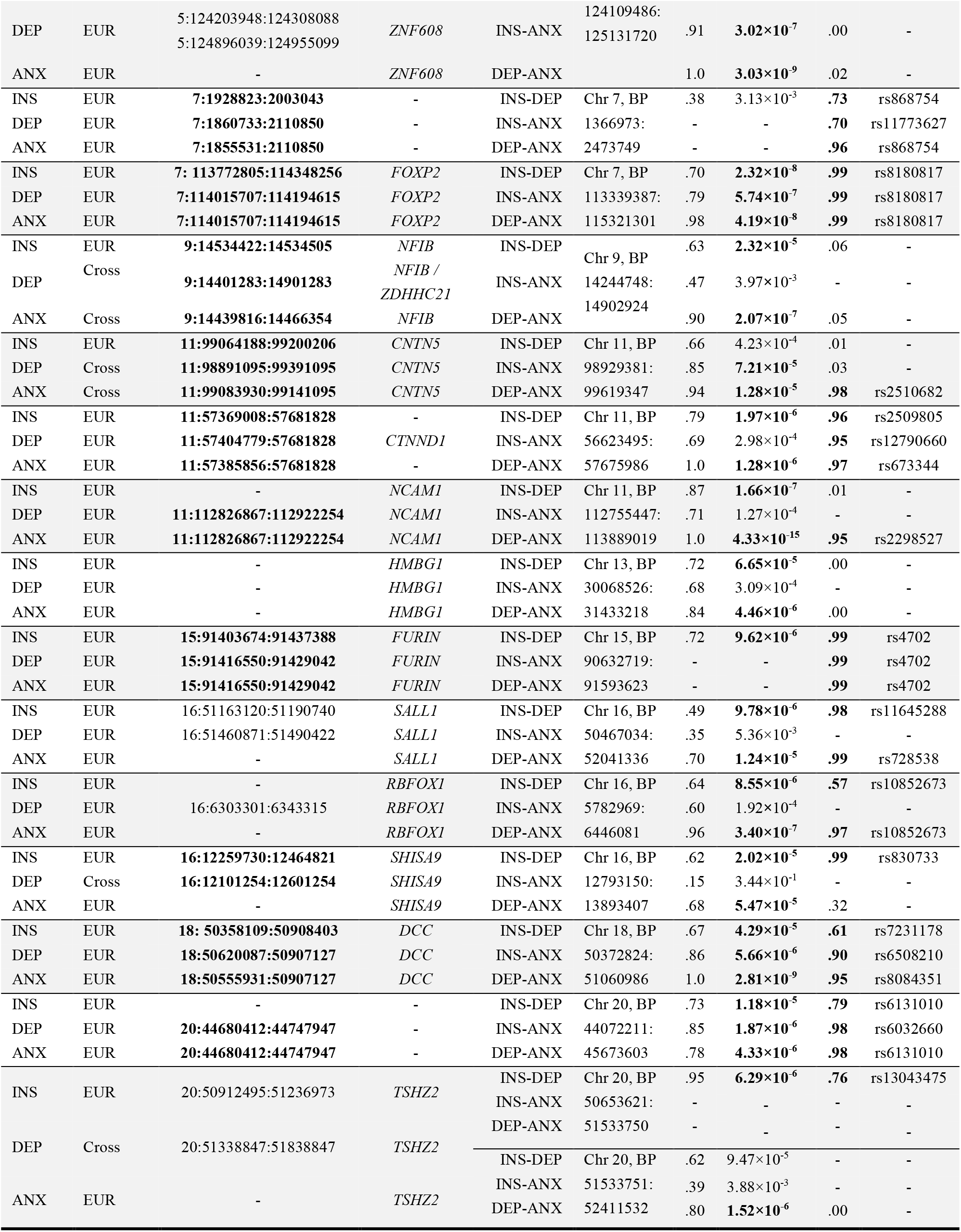
Overview of loci with pleiotropic evidence on the gene or variant level across all three traits. LAVA regions without significant local h2 or loci being unresolved by FLAMES are left blank. H4: posterior probability for the hypothesis that both traits are associated and share a single causal variant. Loci that overlap across traits or metric that pass a certain (e.g. significance) threshold are printed in bold. Without colocalization evidence, we did not perform variant-level mediation analyses.

| Trait | Ancestry | GWS locus<br>(Chr:Start:End) | Gene | Traitpair | Genetic correlation (EUR) |  |  | Colocalization |  |
|---|---|---|---|---|---|---|---|---|---|
|  |  |  |  |  | Region | *r*<sub>g</sub> | *P* | H4 | Top SNP |
| INS | EUR | **1:72512988:74077588** | *NEGR1 / LRRIQ3* | INS-DEP | Chr 1, BP | .88 | **5.17×10<sup>-12</sup>** | .47 | - |
|  |  |  |  | INS-ANX | 72513120: | .85 | **2.20×10<sup>-7</sup>** | .00 | - |
| DEP | EUR | **1:73307570:73991792** | *NEGR1 / LRRIQ3* | DEP-ANX | 73992170 | .97 | **7.31×10<sup>-10</sup>** | .00 | - |
|  |  |  |  | INS-DEP | Chr 1, BP | .56 | **5.56×10<sup>-5</sup>** | .41 | - |
| ANX | EUR | **1:73766431:73991651** | *LRRIQ3* | INS-ANX | 73992171: | .73 | **1.04×10<sup>-6</sup>** | **.77** | rs61767168 |
|  |  |  |  | DEP-ANX | 75132092 | .76 | **2.72×10<sup>-6</sup>** | **.94** | rs7540134 |
| INS | EUR | - | *ARHGAP15* | INS-DEP | Chr 2, BP | .82 | **6.95×10<sup>-5</sup>** | .03 | - |
| DEP | EUR | **2:144146675:144248905** | *ARHGAP15* | INS-ANX | 143259781: | .80 | **5.74×10<sup>-7</sup>** | .01 | - |
| ANX | EUR | **2:144145478:144272229** | *ARHGAP15* | DEP-ANX | 144503688 | 1.0 | **1.06×10<sup>-6</sup>** | **.96** | rs1320139 |
| INS | EUR | 3:71433554:71474216 | *FOXP1* | INS-DEP | Chr 3, BP | .34 | 3.42×10<sup>-3</sup> | - | - |
| DEP | EUR | 3:71748419:71820969 | *FOXP1* | INS-ANX | 71223282: | .75 | **6.04×10<sup>-7</sup>** | .00 | - |
| ANX | EUR | - | *FOXP1* | DEP-ANX | 72334704 | .68 | **7.67×10<sup>-5</sup>** | **.53** | rs1617010 |
| INS | EUR | - | *LPHN3* | INS-DEP | Chr 4, BP | .72 | **6.67×10<sup>-5</sup>** | .01 | - |
| DEP | EUR | - | *LPHN3* | INS-ANX | 61971162: | .41 | 2.20×10<sup>-2</sup> | - | - |
| ANX | EUR | - | *LPHN3* | DEP-ANX | 63337954 | .93 | **1.71×10<sup>-6</sup>** | .01 | - |
| INS | EUR | **5:92995013:93494731** | *NR2F1* | INS-DEP | Chr 5, BP | .51 | 4.96×10<sup>-4</sup> | **.91** | rs72786643 |
| DEP | Cross | **5: 92788598:93288598** | *NR2F1* | INS-ANX | 91956906: | - | - | - | - |
| ANX | EUR | **5:92377803:92487543** | *NR2F1* | DEP-ANX | 93814604 | - | - | **.88** | rs10078721 |
| INS | EUR | - | *ZNF608* | INS-DEP | Chr 5, BP | .69 | **6.47×10<sup>-6</sup>** | .03 | - |
| DEP | EUR | 5:124203948:124308088<br>5:124896039:124955099 | *ZNF608* | INS-ANX | 124109486:<br>125131720 | .91 | **3.02×10<sup>-7</sup>** | .00 | - |
| ANX | EUR | - | *ZNF608* | DEP-ANX |  | 1.0 | **3.03×10<sup>-9</sup>** | .02 | - |
| INS | EUR | **7:1928823:2003043** | - | INS-DEP | Chr 7, BP | .38 | 3.13×10<sup>-3</sup> | **.73** | rs868754 |
| DEP | EUR | **7:1860733:2110850** | - | INS-ANX | 1366973: | - | - | **.70** | rs11773627 |
| ANX | EUR | **7:1855531:2110850** | - | DEP-ANX | 2473749 | - | - | **.96** | rs868754 |
| INS | EUR | **7: 113772805:114348256** | *FOXP2* | INS-DEP | Chr 7, BP | .70 | **2.32×10<sup>-8</sup>** | **.99** | rs8180817 |
| DEP | EUR | **7:114015707:114194615** | *FOXP2* | INS-ANX | 113339387: | .79 | **5.74×10<sup>-7</sup>** | **.99** | rs8180817 |
| ANX | EUR | **7:114015707:114194615** | *FOXP2* | DEP-ANX | 115321301 | .98 | **4.19×10<sup>-8</sup>** | **.99** | rs8180817 |
| INS | EUR | **9:14534422:14534505** | *NFIB* | INS-DEP | Chr 9, BP | .63 | **2.32×10<sup>-5</sup>** | .06 | - |
| DEP | Cross | **9:14401283:14901283** | *NFIB / ZDHHC21* | INS-ANX | 14244748: | .47 | 3.97×10<sup>-3</sup> | - | - |
| ANX | Cross | **9:14439816:14466354** | *NFIB* | DEP-ANX | 14902924 | .90 | **2.07×10<sup>-7</sup>** | .05 | - |
| INS | EUR | **11:99064188:99200206** | *CNTN5* | INS-DEP | Chr 11, BP | .66 | 4.23×10<sup>-4</sup> | .01 | - |
| DEP | Cross | **11:98891095:99391095** | *CNTN5* | INS-ANX | 98929381: | .85 | **7.21×10<sup>-5</sup>** | .03 | - |
| ANX | Cross | **11:99083930:99141095** | *CNTN5* | DEP-ANX | 99619347 | .94 | **1.28×10<sup>-5</sup>** | **.98** | rs2510682 |
| INS | EUR | **11:57369008:57681828** | - | INS-DEP | Chr 11, BP | .79 | **1.97×10<sup>-6</sup>** | **.96** | rs2509805 |
| DEP | EUR | **11:57404779:57681828** | *CTNND1* | INS-ANX | 56623495: | .69 | 2.98×10<sup>-4</sup> | **.95** | rs12790660 |
| ANX | EUR | **11:57385856:57681828** | - | DEP-ANX | 57675986 | 1.0 | **1.28×10<sup>-6</sup>** | **.97** | rs673344 |
| INS | EUR | - | *NCAM1* | INS-DEP | Chr 11, BP | .87 | **1.66×10<sup>-7</sup>** | .01 | - |
| DEP | EUR | **11:112826867:112922254** | *NCAM1* | INS-ANX | 112755447: | .71 | 1.27×10<sup>-4</sup> | - | - |
| ANX | EUR | **11:112826867:112922254** | *NCAM1* | DEP-ANX | 113889019 | 1.0 | **4.33×10<sup>-15</sup>** | **.95** | rs2298527 |
| INS | EUR | - | *HMBG1* | INS-DEP | Chr 13, BP | .72 | **6.65×10<sup>-5</sup>** | .00 | - |
| DEP | EUR | - | *HMBG1* | INS-ANX | 30068526: | .68 | 3.09×10<sup>-4</sup> | - | - |
| ANX | EUR | - | *HMBG1* | DEP-ANX | 31433218 | .84 | **4.46×10<sup>-6</sup>** | .00 | - |
| INS | EUR | **15:91403674:91437388** | *FURIN* | INS-DEP | Chr 15, BP | .72 | **9.62×10<sup>-6</sup>** | **.99** | rs4702 |
| DEP | EUR | **15:91416550:91429042** | *FURIN* | INS-ANX | 90632719: | - | - | **.99** | rs4702 |
| ANX | EUR | **15:91416550:91429042** | *FURIN* | DEP-ANX | 91593623 | - | - | **.99** | rs4702 |
| INS | EUR | 16:51163120:51190740 | *SALL1* | INS-DEP | Chr 16, BP | .49 | **9.78×10<sup>-6</sup>** | **.98** | rs11645288 |
| DEP | EUR | 16:51460871:51490422 | *SALL1* | INS-ANX | 50467034: | .35 | 5.36×10<sup>-3</sup> | - | - |
| ANX | EUR | - | *SALL1* | DEP-ANX | 52041336 | .70 | **1.24×10<sup>-5</sup>** | **.99** | rs728538 |
| INS | EUR | - | *RBFOX1* | INS-DEP | Chr 16, BP | .64 | **8.55×10<sup>-6</sup>** | **.57** | rs10852673 |
| DEP | EUR | 16:6303301:6343315 | *RBFOX1* | INS-ANX | 5782969: | .60 | 1.92×10<sup>-4</sup> | - | - |
| ANX | EUR | - | *RBFOX1* | DEP-ANX | 6446081 | .96 | **3.40×10<sup>-7</sup>** | **.97** | rs10852673 |
| INS | EUR | **16:12259730:12464821** | *SHISA9* | INS-DEP | Chr 16, BP | .62 | **2.02×10<sup>-5</sup>** | **.99** | rs830733 |
| DEP | Cross | **16:12101254:12601254** | *SHISA9* | INS-ANX | 12793150: | .15 | 3.44×10<sup>-1</sup> | - | - |
| ANX | EUR | - | *SHISA9* | DEP-ANX | 13893407 | .68 | **5.47×10<sup>-5</sup>** | .32 | - |
| INS | EUR | **18: 50358109:50908403** | *DCC* | INS-DEP | Chr 18, BP | .67 | **4.29×10<sup>-5</sup>** | **.61** | rs7231178 |
| DEP | EUR | **18:50620087:50907127** | *DCC* | INS-ANX | 50372824: | .86 | **5.66×10<sup>-6</sup>** | **.90** | rs6508210 |
| ANX | EUR | **18:50555931:50907127** | *DCC* | DEP-ANX | 51060986 | 1.0 | **2.81×10<sup>-9</sup>** | **.95** | rs8084351 |
| INS | EUR | - | - | INS-DEP | Chr 20, BP | .73 | **1.18×10<sup>-5</sup>** | **.79** | rs6131010 |
| DEP | EUR | **20:44680412:44747947** | - | INS-ANX | 44072211: | .85 | **1.87×10<sup>-6</sup>** | **.98** | rs6032660 |
| ANX | EUR | **20:44680412:44747947** | - | DEP-ANX | 45673603 | .78 | **4.33×10<sup>-6</sup>** | **.98** | rs6131010 |
| INS | EUR | 20:50912495:51236973 | *TSHZ2* | INS-DEP | Chr 20, BP | .95 | **6.29×10<sup>-6</sup>** | **.76** | rs13043475 |
|  |  |  |  | INS-ANX | 50653621: | - | - | - | - |
| DEP | Cross | 20:51338847:51838847 | *TSHZ2* | DEP-ANX | 51533750 | - | - | - | - |
|  |  |  |  | INS-DEP | Chr 20, BP | .62 | 9.47×10<sup>-5</sup> | - | - |
| ANX | EUR | - | *TSHZ2* | INS-ANX | 51533751: | .39 | 3.88×10<sup>-3</sup> | - | - |
|  |  |  |  | DEP-ANX | 52411532 | .80 | **1.52×10<sup>-6</sup>** | .00 | - |

### Trivariate genetic overlap on the genome-wide level

Combined genome-wide analysis of all three phenotypes in a trivariate approach has not been performed previously, but has the potential to distinguish the portion of genetic overlap shared between all three conditions, and the proportion of genetic overlap unique to each pair of conditions^37^. We therefore modeled genome-wide genetic overlap across three traits simultaneously using Trivariate MiXeR^37^, a recently developed extension of the established univariate and bivariate MiXeR models^39,50^. These results deepen findings from standard pairwise genetic correlations (*r*_g_) estimated with LD Score regression^51^ (Supplementary Note 3).

We observed a genetic overlap of 55% across all three traits (Figure 1). The genetic signal of INS overlapping with DEP and with ANX was of comparable size (78% and 79% respectively) and concerned the same SNPs, since 98% of DEP overlapped with ANX (Supplementary Table 2). However, the correlation of effect sizes within the overlap was stronger between INS-ANX (*r*_g-shared_=.85, SD=.09) than between INS-DEP (*r*_g-shared_=.63, SD=.04). As expected based on previous studies^17,20,23,24^, the strongest correlation of effect sizes was observed for the overlapping SNPs between DEP-ANX (*r*_g-shared_=.94, SD=.04). Notably, 21% of INS variants were not shared with ANX or DEP. While ANX also contained a unique genetic component, this was accompanied by larger polygenicity (number of non-null SNPs; *π*_ANX_=14,463; *π*_DEP_=12,474, *π*_INS_=12,180; Supplementary Table 2), which could be driven by the pooling of various anxiety disorders included in the ANX GWAS.

**Figure 1.**
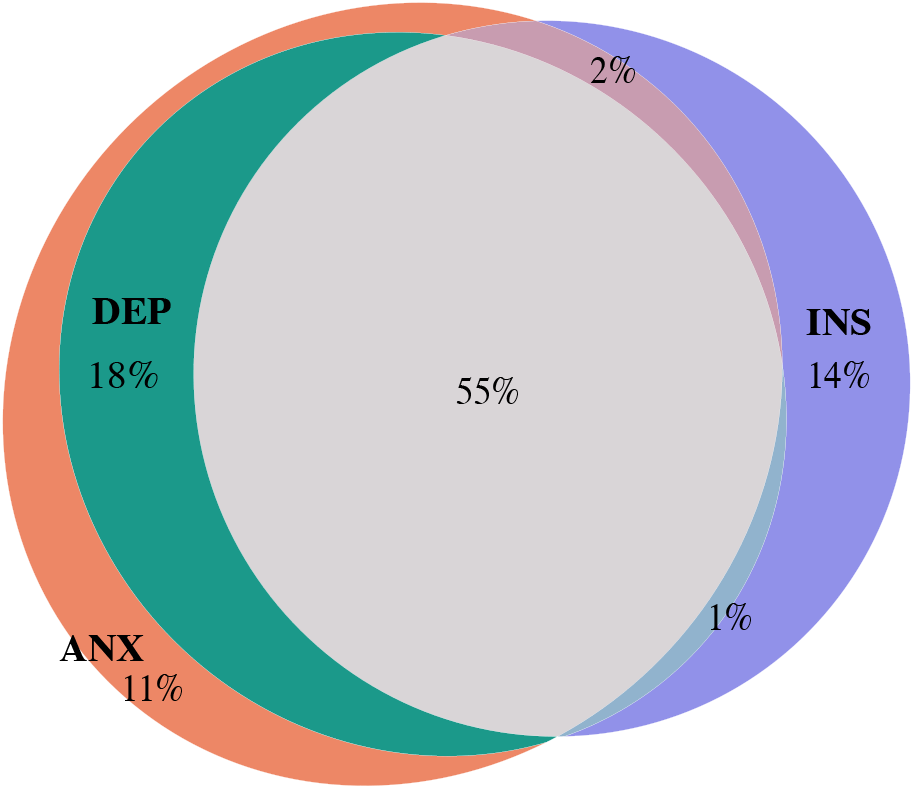
Genome-wide genetic overlap of INS, ANX and DEP. a) Trivariate overlap estimated with MiXeR (Supplementary Table 2). Each circle (i.e. trait) has a size proportional to the number of trait-influencing variants (i.e. polygenicity) and overlapping areas proportional to the number of variants shared among respective traits. Percentages in the diagram are given with respect to the combined total area of all three traits (and sum to 101% due to rounding).

### Identifying pleiotropic loci across conditions

We aimed to move beyond global patterns and identify regions in the genome (i.e. loci) with shared genetic risk. First, we investigated which of the loci that reached genome-wide significance (GWS; *p*<5×10^−8^) in one trait physically overlapped with GWS loci in the other traits. The EUR GWAS summary statistics included 40, 235, and 248 GWS loci for ANX^17^, DEP^20^ and INS^16^ respectively. Cross-ancestry GWAS summary statistics of ANX^17^ and DEP^19^ included 10 and 47 genome-wide significant loci, respectively, that were not identified in the EUR-only analyses. One locus was GWS in the AFR summary statistics for ANX^17^, none for DEP^19^. This totalled 580 GWS loci in our three traits across these ancestries. Thirty-two percent of these loci were overlapping with at least one locus of another phenotype in the triad, spanning 87 unique locations in the genome. Of all EUR and cross-ancestry loci (Supplementary Table 3), nine overlapped across all three phenotypes, 49 physically overlapped between INS-DEP, 26 between DEP-ANX, and three between INS-ANX (Figure 2; Supplementary Table 4). The GWS locus of ANX in the AFR sample did not overlap with any locus of DEP or INS.

**Figure 2.**
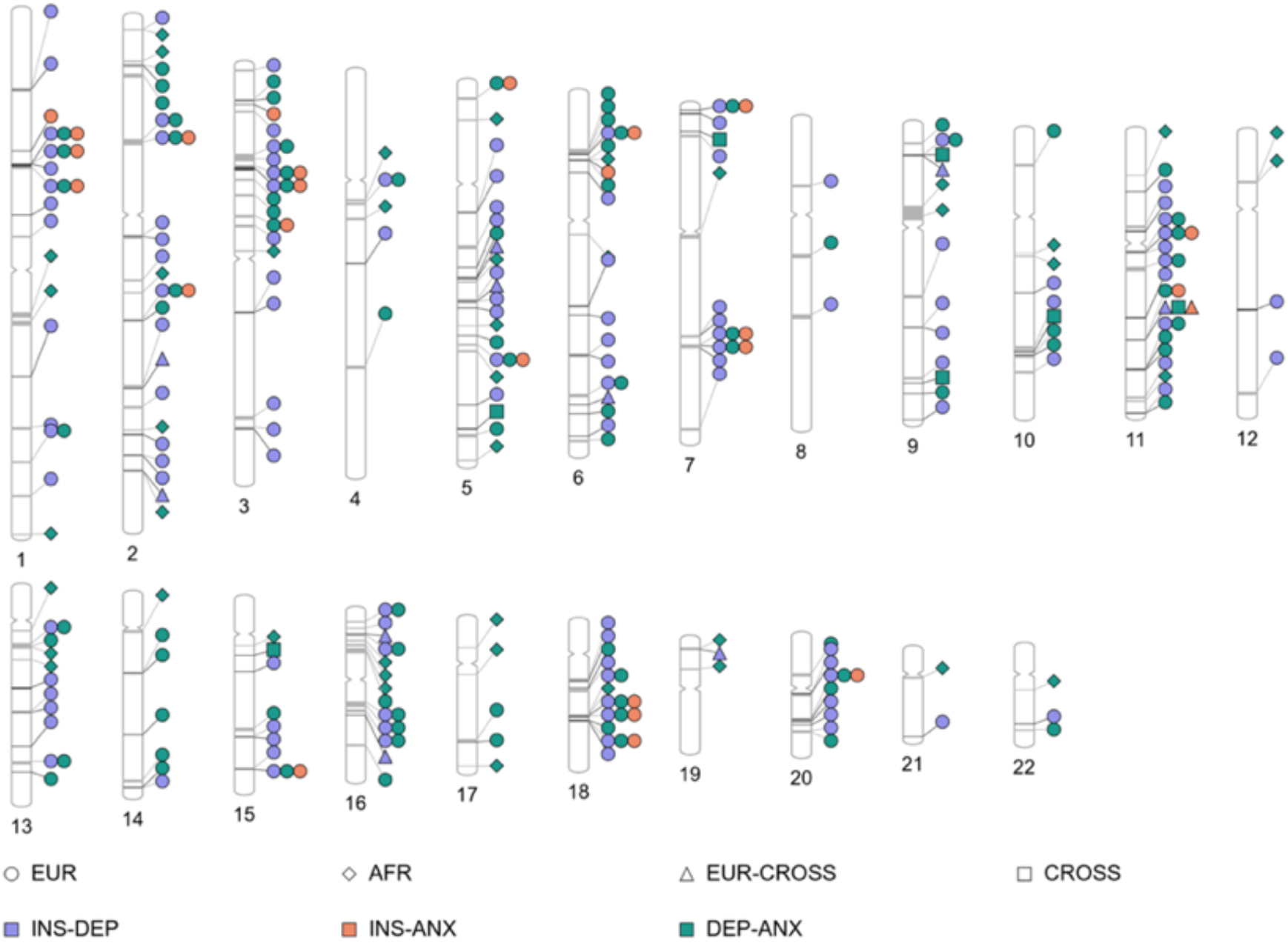
Loci with shared genetic risk across ANX, DEP and INS. Ideogram with each chromosome showing the location of genome-wide significant loci (dark grey lines) that were physically overlapping across INS, DEP and ANX GWAS summary statistics (both EUR or cross-ancestry, or EUR for INS and cross-ancestry for DEP/ANX). Markers connected by light grey lines had significant correlating GWAS signal across trait pairs (i.e. identified through LAVA).

Second, we identified regions with significant cross-disorder local genetic correlations by using LAVA^41^ on the EUR summary statistics. We first screened *n*=2,495 non-overlapping genomic regions for sufficient univariate genetic signal in each trait (*n*=582 survived *p*_univ_<1×10^−4^) before bivariate genetic correlations between INS-ANX, INS-DEP, and/or ANX-DEP were estimated. Local *r*_g_ estimates were significant after correcting for multiple testing (*p*_bivar_<.05/582=8.59×10^−5^) in ten loci for all three trait pairs (Supplementary Table 5-6). Seventeen loci were significant for INS-DEP as well as DEP-ANX, and 3 loci showed significant local *r*_g_ between both INS-ANX and DEP-ANX. Most loci were only significant for one trait pair (*n*_INS-DEP_=46, *n*_INS-ANX_=5, *n*_DEP-ANX_=27).

Of all 108 unique LAVA-identified loci, *n*=39 loci physically overlapped with any shared GWS locus described above. These LAVA regions were almost always identified for the same (and an additional) trait pair as the GWS locus (Supplementary Table 6). In total, 195 loci were identified to be shared across at least one trait pair (*n*_GWS_=87, *n*_LAVA_=108), which could be reduced to 146 unique locations given the overlap across approaches. We continued our downstream analyses with all 195 loci with cross-disorder evidence.

### Do shared loci also share the same putative-causal variant(s)?

We tested each of the cross-disorder 195 loci identified above for having an observed genetic signal consistent with a shared causal variant. For this purpose, we ran colocalization analysis with COLOC^33^ on the GWS and LAVA loci for each relevant trait pair. These loci colocalized to the same variant in 50% of locus pairs (Supplementary Table 7). Of these loci, still 85.7% colocalize in a sensitivity analysis relaxing the single-variant assumption, despite the best available reference panel not being a perfect match and significantly smaller than the meta-analyses (methods). Thirteen loci showed colocalization evidence across all three traits (Table 1), of which five loci were also assigned the same top SNP with the highest posterior probability (PP) of being the most likely-shared causal variant across traits. See Supplementary Note 4 for colocalization results specified by locus type (i.e. GWS *vs* LAVA).

### Shared INS-DEP-ANX loci converge on the gene level

We further explored shared likely-causal effects in cross-disorder loci by assessing the level of pleiotropy on the gene-level. We ran FLAMES on all overlapping GWS and LAVA loci, and found 84 effector genes predicted for at least two traits. Seventeen genes were predicted to be effector genes for all three conditions (Table 1), *n*=38 were uniquely shared between DEP-INS, *n*=2 were uniquely shared between ANX-INS, and *n*=27 were uniquely shared between ANX-DEP (Figure 3; Supplementary Table 8). Out of all predicted effector genes, 60% were predicted for at least two traits (81% ANX, 80% INS, 71% DEP). However, the remaining 40% includes loci where FLAMES resolved its prediction for only one trait and could not predict an effector gene with certainty for the other(s), which dilutes estimates of concordance. When considering shared loci where at least two genes were prioritized, these genes are concordant in 80% of overlapping loci. These results show that in the majority of FLAMES resolved loci of ANX, DEP and INS, the GWAS signal converges onto the same predicted effector genes.

**Figure 3.**
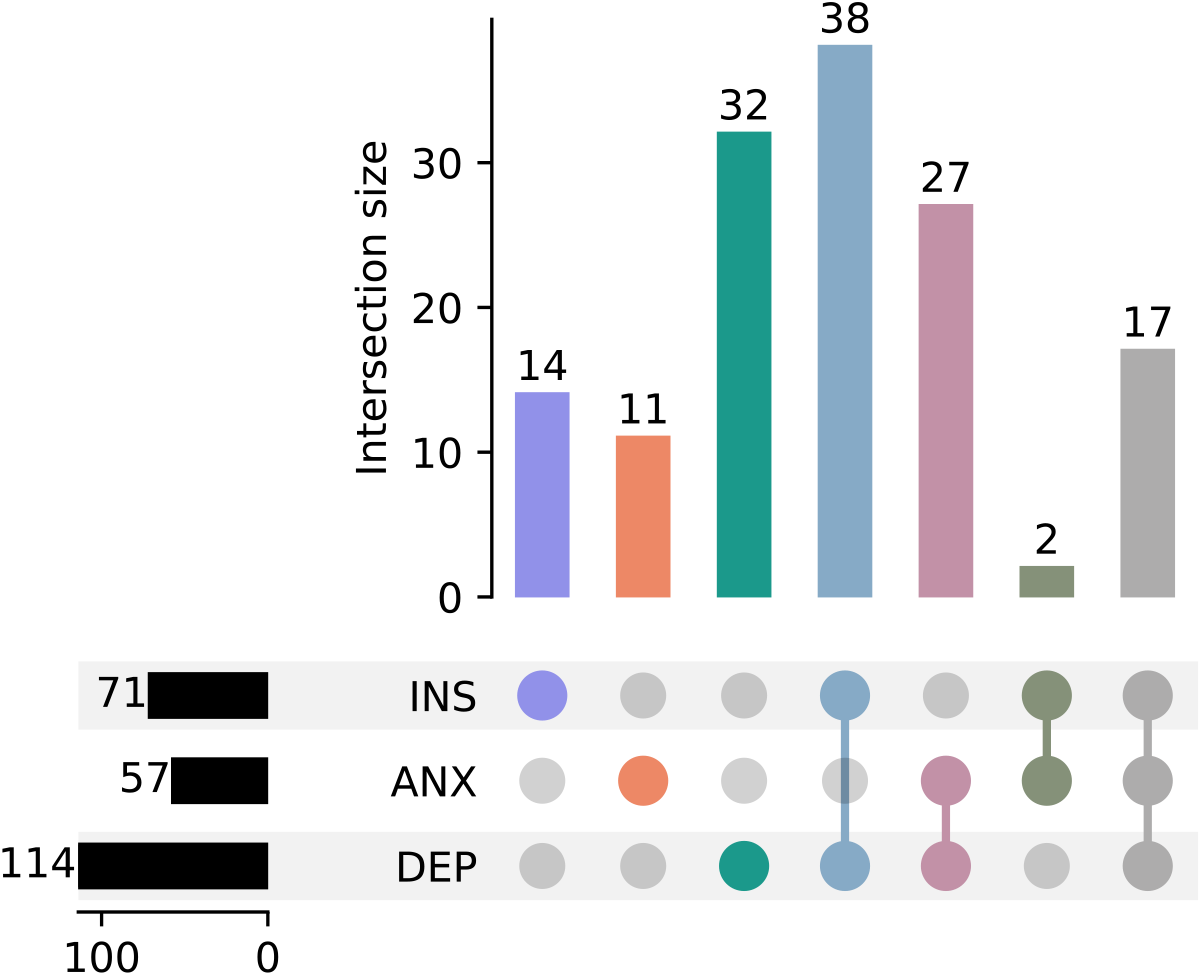
Number of overlapping effector genes across conditions. We aggregated the identified effector genes (by FLAMES) across ancestries and across traits to give insight into the number of shared genes within each of these domains.

### Biological mechanisms of shared effector genes

We performed Gene Ontology (GO)^48^ enrichment analysis on three distinct sets of shared predicted effector genes (*n*_INS-DEP-ANX_=17, *n*_DEP-ANX_=27, *n*_INS-DEP_=38). Enrichment analyses in tissue and cell-type gene expression markers are presented in Supplementary Note 6. We found significantly associated biological pathways and cellular components with all three sets (Figure 4, Supplementary Table 9), many of which are strongly related to neurological development and synapse function despite testing for enrichment in the broad array of gene-sets. The function of the two predicted effector genes uniquely shared between ANX-INS (*PDE4B, FTO*) were aligned with the enriched gene-sets (Supplementary Note 5).

**Figure 4.**
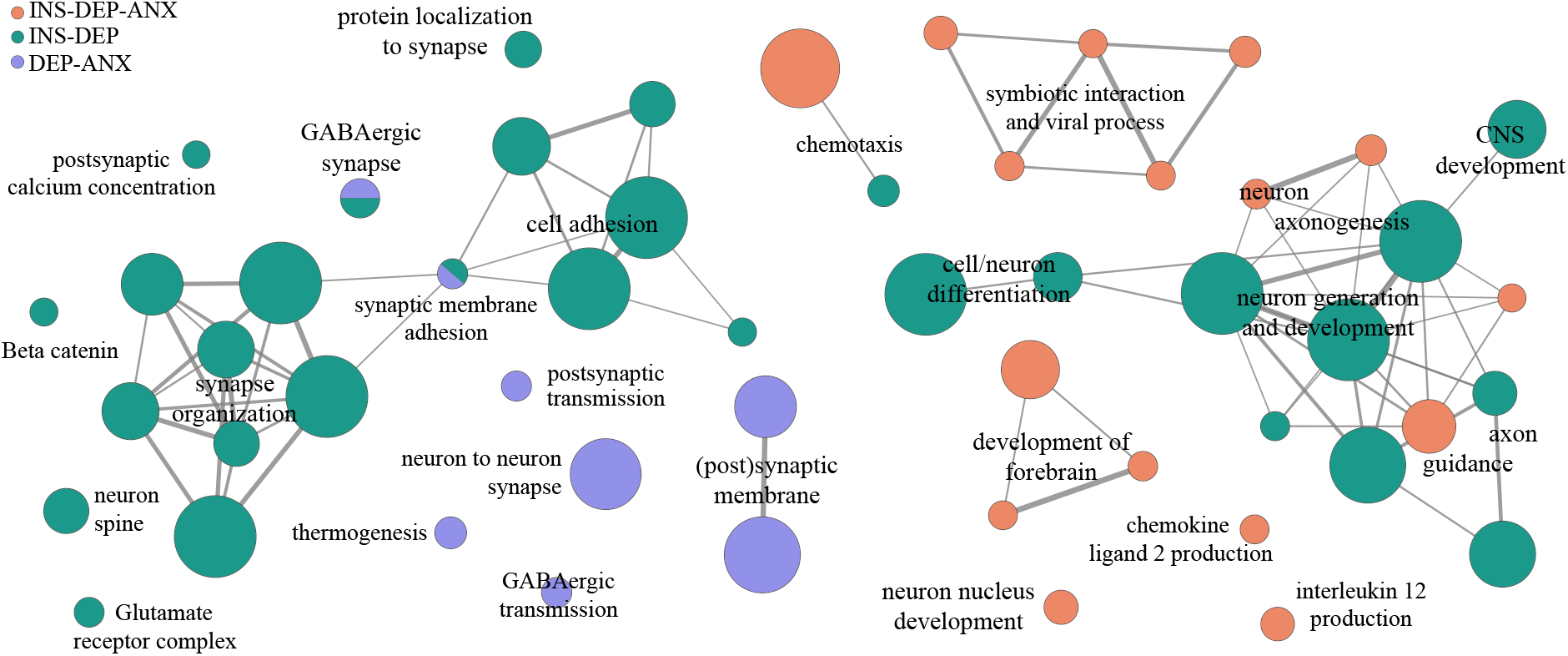
Biological processes and cellular component enriched for true pleiotropic genes. Predicted effector genes that were shared among two or three traits were significantly enriched (*p*<1.90×10^−6^) in Gene Ontology (GO) biological processes and cellular components. Note that only one gene was uniquely shared between INS and ANX, and was therefore not tested. Graph was made using Cytoscape^57^ and EnrichmentMap^65^. Pathway similarity scores (Jaccard and overlap combined coefficient=.5) were used as edges between the gene-sets (see the Nature Protocol by Reimand et al.^59^ for further details)

Despite having no genes in common, the predicted effector genes uniquely shared by ANX-DEP and INS-DEP were enriched in the same gene-sets (Cellular Component GABAergic synapse; Biological Processes synaptic membrane adhesion), providing evidence for pathway-level convergence. Predicted effector genes uniquely shared between INS-DEP were enriched in processes related to cell adhesion and synaptic structuring, neuronal development and differentiation, synaptic signaling and plasticity. The predicted effector genes uniquely shared between ANX-DEP were enriched in the (post)synaptic membrane, postsynaptic modulation of synaptic transmission and the regulation of GABAergic neurotransmission.

Genes with pleiotropic evidence across all three conditions were enriched in processes that are very typically found across psychiatric genetic literature^52^, such as axon guidance, and forebrain development. This may reflect more general neurodevelopmental liability rather than mechanisms specifically shared between insomnia, depression and anxiety disorders. This is reflected in the enrichment for general brain tissue and neuronal cell-type enrichments for the shared gene sets (Supplementary Note 6). Additionally we find that processes related to viral infection and inflammation, aligning with prior observations^53^. The genes involved in chemokine 2 and interleukin 12 production and viral processes (*FOXP1, HMGB1, FURIN* and *NCAM1)* that are shared between INS-DEP-ANX can indicate immune processes, but are also critical for neurodevelopment, growth and neuronal signalling^54–56^.

### Shared effector genes are drug targets for INS, DEP ANX through synaptic membrane mechanisms

We used the OpenTargets database^60^ to explore if any of our 84 shared predicted effector genes have been investigated as potential drug targets for INS, DEP or ANX. We found that nine (10.7%) of our genes are targets of approved or currently investigated drugs. This is comparable with the fraction (7.5%) of genes that are in OpenTargets as a known drug target (1,518 genes) from those that FLAMES could prioritize (20,260 genes). Interestingly, six of these nine drug targets have previously been investigated as targets of 43 drugs for INS, DEP and/or ANX (Supplementary Table 11). All six (*CACNA2D3, DRD2, ESR1, GRIA1, GRM5* and *PDE4B*) were identified as shared predicted effectors for trait pairs (*n*_DEP-ANX_=3, *n*_INS-DEP_=2, *n*_INS-ANX_=1) rather than for the INS-DEP-ANX triad. Five out of these six genes (all except *ESR1*) were also part of our enriched gene-sets involved in the (post)synaptic membrane, neuron-to-neuron synapse and neuron spine. Finding five out of nine drugs to target synaptic genes is a significant enrichment (*p* = 0.04, hypergeometric test), with only 2.18 expected synaptic genes expected for nine randomly selected drug targets. The fraction of synaptic genes is similar in the drug database (24.2%) and the predicted effector genes (29.1%). Other significantly enriched pathways or cellular components are likely only minimally druggable through the genes we identified, since these GO-terms contained only one or two druggable genes.

Almost all of the 43 drugs are approved or in late-stage clinical trials for ANX (*n*=17), DEP (*n*=13) or both (*n*=12; Supplementary Table 11). The majority of these drugs (*n*=35) are antagonists for dopamine receptor D2, or glutamate receptor 1 or 5 (targeting *DRD2, GRIA1* or *GRM5* respectively)^49^. Two approved drugs for DEP that target *DRD2* (trimiparine and quetiapine) are also prescribed off-label for INS, and for ANX and INS, respectively^49^. The only drug with a sole indication for INS was atagabalin, an inhibitor of voltage-dependent calcium channel α2δ3 (target gene *CACNA2D3*) reaching phase 2 clinical trials.

Overall, we find that a significant subset of the pathways we identified to be enriched for predicted effector genes shared between INS, DEP and ANX show promise for continued drug development, with many investigated and approved drugs that target a shared predicted effector gene having effects on multiple of these disorders simultaneously.

## Discussion

Insomnia, depression and anxiety disorders frequently co-occur and share a substantial proportion of their genetic risk^12^. The current study combines existing methods into a hierarchical framework, tracing down genome-wide overlap to variant level effects. On the highest level, we find 55% trivariate genetic overlap with high correlations of effect sizes of overlapping variants. We subsequently identify 195 genomic loci with statistical evidence of shared genetic risk across (pairs of) traits. The majority of these loci contain a shared putative-causal variant and/or shared predicted effector gene. Predicted effector genes shared between different trait pairs converge on processes related to the GABAergic synapse and synaptic membrane adhesion, and include targets of twelve previously investigated and/or approved medications with an indication for at least two of the ANX, DEP, INS traits. Altogether, these findings prioritize molecular genetic liability factors shared across insomnia, depression and anxiety disorders and highlight transdiagnostic biological mechanisms that are potentially therapeutically actionable.

Previous work has suggested that subsets of the same 10,000-15,000 variants explain most of the SNP heritability of major psychiatric disorders, with genetic differences between conditions arising from different directions and magnitudes of effect of these variants^20^. Our Trivariate MiXeR analysis shows that, although insomnia shares a substantial proportion of its genetic architecture with depression and anxiety disorders (as is in line with pairwise previous studies^21,24,27^), about one fifth of its genetic signal appears distinct. Since previous results showed that insomnia loci colocalize with loci from traits clustering into a psychiatric and metabolic component^61^, future work could investigate whether the genetic signal that appears to be unique to insomnia in this study, overlaps with the genetics of metabolic traits. This may help to disentangle the genetic heterogeneity of insomnia and inform stratified therapeutic approaches.

By applying the multi-level analysis framework proposed here, we demonstrated that the majority of the shared genetic risk loci consistently points to the same putative-causal variant and gene. Specifically, we find that approximately two thirds of loci, when containing enough genetic signal, colocalize between these three traits. Predicted effector genes appear to be shared in approximately three quarters of the investigated loci. In some loci, different effector genes were prioritized in the presence of evidence for a shared causal variant (Supplementary Note 7). This underscores that although overlapping GWAS signal between insomnia, depression and anxiety disorders can generally be considered non-spurious pleiotropic, the loci we highlight should be considered a prioritized set of hypotheses for laboratory validation, rather than definitive evidence of shared biology^62^.

The majority of the druggable genes that we identified as shared predicted effector genes across insomnia, depression and anxiety disorders have previously been investigated as drug targets for these conditions. Off-label prescriptions that are used transdiagnostically indicate promise for continued development of therapeutics targeting genes in e.g. GABA-ergic and glutamatergic signalling in the synapse. Of course, the neurodevelopmental processes underlying predispositions for ANX, DEP and INS generally precede diagnosis, yet altering synapse organization and plasticity in adulthood might still result in therapeutic benefits as suggested by these results The findings of this study should be considered within the context of a small total explained variance of the genetic component investigated: we focus only on the genetic signal shared between traits, that individually have very modest SNP-based heritability. Nevertheless, the relevance of the biological processes we identify is supported by previous pharmacological^63–65^ and neurobiological^66–69^ evidence. For example, the shared predicted effector genes enriched in the GABAergic neuron, synaptic adhesion and the glutamatergic receptor complex gene sets are mostly involved in the regulation of tans-synaptic adhesion, receptor anchoring and signaling modulation, all of which fine-tune synaptic plasticity mechanisms^70–76^. Disturbances of synaptic plasticity have previously been reported in patients suffering from anxiety disorders, depression and insomnia^77,78^. Our findings extend previous literature by showing that genes in these pathways are impacted by common genetic risk variants across conditions.

Important limitations of this work include a source of potential inflation in the observed genetic overlap cannot be accounted for in this study, that is phenotype conflation. As mentioned above, the initial insomnia, depression, and anxiety disorders GWAS analyses could have been biased by a level of misdiagnosis, unrecognized comorbidity resulting from ascertainment bias^79^, or the use of both unscreened and supernatural controls^80^. Although our sensitivity analyses showed that the genetic correlations were robust to UKB sample overlap and broad phenotype definitions (Supplementary Note 1), we cannot rule out symptom overlap across diagnoses and other cohorts contributing participants to more than one trait. The observed overlap should therefore be interpreted as shared genetic liability of co-occurring phenotypes rather than as evidence of shared biology independent of co-occurrence. Future studies may address these potential sources of spurious pleiotropy by careful longitudinal assessment, and the selection of control subjects screened to only exclude the target conditions^81^. Similarly, underestimation of true genetic overlap can occur when the original GWAS summary statistics lack sufficient statistical power to detect genome-wide significant loci or significant local heritability. The GWAS of anxiety disorders was the least powered GWAS used in our study, which likely contributed to e.g. the smaller number of overlapping predicted effector genes observed, which should not be interpreted as evidence of no overlap. In general, it is possible that loci, genes or variants solely shared between a trait pair in this study will become shared across the full triad when GWAS power further increases.

In conclusion, this study confirms a genetically interconnected basis for insomnia, depression and anxiety disorders across multiple levels of investigation. We provide a shortlist of pleiotropic variants, genes and pathways, offering potential candidates for future pathway-tailored therapeutic targets with transdiagnostic benefits.

## Data availability

This study relies on previously published GWAS summary statistics as cited in the main text. Access to 23andMe summary statistics requires application via https://research.23andme.com/dataset-access/. The data generated in the current manuscript is available in the Supplementary Tables.

## Code availability

The code to obtain the results presented in this manuscript will be available via https://github.com/EPTissink/ upon publication.

## Acknowledgements

This work has received funding from ZonMw, The Hague, The Netherlands, project 09120011910032 REMOVE, and from the European Union: the European Research Council (ERC), Brussels, Belgium, Advanced Grant 101055383 OVERNIGHT (E.T. and E.J.W.V.S.). M.S. was funded by NWO Gravitation: BRAINSCAPES: A Roadmap from Neurogenetics to Neurobiology (Grant No. 024.004.012). Views and opinions expressed are however those of the author(s) only and do not necessarily reflect those of the European Union or the European Research Council Executive Agency. Neither the European Union nor the granting authority can be held responsible for them. The funders had no role in the design, data collection, data analysis, and reporting of this study. We would like to thank Marie Verbanck and Martin Tournaire for insightful discussions. We would like to thank the research participants and employees of 23andMe Research Institute for making this work possible. Earlier versions of this report were included in the doctoral thesis of M.S. and preprinted on *medRxiv*.

## Author Contributions

M.S.: Formal Analysis, Visualization, Writing – Original. A.A.S.: Software, Writing – Review & Editing. C.R.: Software, Writing – Review & Editing. E.F.: Resources, Writing – Review & Editing. R.P.: Resources; Writing – Review & Editing. D.P.: Conceptualization, Writing – Review & Editing. E.J.W.v.S.: Conceptualization, Funding Acquisition, Writing – Review & Editing. E.T.: Conceptualization, Data Curation, Formal Analysis, Supervision, Visualization, Writing – Original.

## References

1. American Psychiatric Association. Diagnostic and Statistical Manual of Mental Disorders, Fifth Edition. (DSM-5). https://cir.nii.ac.jp/crid/1573950399819987840 (2013).

2. Soehner, A. M. & Harvey, A. G. Prevalence and Functional Consequences of Severe Insomnia Symptoms in Mood and Anxiety Disorders: Results from a Nationally Representative Sample. Sleep 35, 1367–1375 (2012).

3. Ohayon, M. M. & Roth, T. Place of chronic insomnia in the course of depressive and anxiety disorders. J. Psychiatr. Res. 37, 9–15 (2003).

4. Roth, T. et al. Sleep Problems, Comorbid Mental Disorders, and Role Functioning in the National Comorbidity Survey Replication. Biol. Psychiatry 60, 1364–1371 (2006).

5. Leerssen, J. et al. Treating Insomnia with High Risk of Depression Using Therapist-Guided Digital Cognitive, Behavioral, and Circadian Rhythm Support Interventions to Prevent Worsening of Depressive Symptoms: A Randomized Controlled Trial. Psychother. Psychosom. 91, 168–179 (2022).

6. de Lange, S. C. et al. Multimodal brain imaging of insomnia, depression and anxiety symptoms indicates transdiagnostic commonalities and differences. Nat. Ment. Health 3, 517–529 (2025).

7. Neale, M. C. & Kendler, K. S. Models of comorbidity for multifactorial disorders. Am. J. Hum. Genet. 57, 935–953 (1995).

8. Staner, L. Comorbidity of insomnia and depression. Sleep Med. Rev. 14, 35–46 (2010).

9. Ruberto, V. L., Jha, M. K. & Murrough, J. W. Pharmacological Treatments for Patients with Treatment-Resistant Depression. Pharmaceuticals 13, 116 (2020).

10. Ansara, E. D. Management of treatment-resistant generalized anxiety disorder. Ment. Health Clin. 10, 326–334 (2020).

11. Minikel, E. V., Painter, J. L., Dong, C. C. & Nelson, M. R. Refining the impact of genetic evidence on clinical success. Nature 629, 624–629 (2024).

12. Lind, M. J. et al. An examination of the etiologic overlap between the genetic and environmental influences on insomnia and common psychopathology. Depress. Anxiety 34, 453–462 (2017).

13. Madrid-Valero, J. J., Rubio-Aparicio, M., Gregory, A. M., Sánchez-Meca, J. & Ordoñana, J. R. The heritability of insomnia: Systematic review and meta-analysis of twin studies. Sleep Med. Rev. 58, 101437 (2021).

14. Sullivan, P. F., Michael Neale, F. C. & Kendler, K. S. Reviews and Overviews Genetic Epidemiology of Major Depression: Review and Meta-Analysis. Am J Psychiatry vol. 15710 http://views.vcu.edu/pub/mx/examples/mdreview. (2000).

15. Hettema, J. M., Neale, M. C. & Kendler, K. S. A Review and Meta-Analysis of the Genetic Epidemiology of Anxiety Disorders. Am. J. Psychiatry 158, 1568–1578 (2001).

16. Jansen, P. R. et al. Genome-wide analysis of insomnia in 1,331,010 individuals identifies new risk loci and functional pathways. Nat. Genet. 51, 394–403 (2019).

17. Friligkou, E. et al. Gene discovery and biological insights into anxiety disorders from a large-scale multi-ancestry genome-wide association study. Nat. Genet. 56, 2036–2045 (2024).

18. Strom, N. I. et al. Genome-wide association study of major anxiety disorders in 122,341 European-ancestry cases identifies 58 loci and highlights GABAergic signaling. 2024.07.03.24309466 Preprint at 10.1101/2024.07.03.24309466 (2024).

19. Meng, X. et al. Multi-ancestry genome-wide association study of major depression aids locus discovery, fine mapping, gene prioritization and causal inference. Nat. Genet. 56, 222–233 (2024).

20. Als, T. D. et al. Depression pathophysiology, risk prediction of recurrence and comorbid psychiatric disorders using genome-wide analyses. Nat. Med. 29, 1832–1844 (2023).

21. Romero, C. et al. Exploring the genetic overlap between twelve psychiatric disorders. Nat. Genet. 54, 1795–1802 (2022).

22. Wray, N. R., Lee, S. H. & Kendler, K. S. Impact of diagnostic misclassification on estimation of genetic correlations using genome-wide genotypes. Eur. J. Hum. Genet. 20, 668–674 (2012).

23. Grotzinger, A. D. et al. Genetic architecture of 11 major psychiatric disorders at biobehavioral, functional genomic and molecular genetic levels of analysis. Nat. Genet. 54, 548–559 (2022).

24. Tesfaye, M. et al. Identification of Novel Genomic Loci for Anxiety and Extensive Genetic Overlap with Psychiatric Disorders. medRxiv 2023.09.01.23294920 (2024) doi:10.1101/2023.09.01.23294920.

25. The Brainstorm Consortium et al. Analysis of shared heritability in common disorders of the brain. Science 360, eaap8757 (2018).

26. Lee, P. H. et al. Genomic Relationships, Novel Loci, and Pleiotropic Mechanisms across Eight Psychiatric Disorders. Cell 179, 1469–1482.e11 (2019).

27. O’Connell, K. S. et al. Characterizing the Genetic Overlap Between Psychiatric Disorders and Sleep-Related Phenotypes. Biol. Psychiatry 90, 621–631 (2021).

28. Baranova, A., Cao, H. & Zhang, F. Shared genetic liability and causal effects between major depressive disorder and insomnia. Hum. Mol. Genet. 31, 1336–1345 (2022).

29. Fisher, R. A. The Correlation between Relatives on the Supposition of Mendelian Inheritance. Trans. R. Soc. Edinb. 52, 399–433 (1919).

30. Falconer, D. S. Introduction to Quantitative Genetics. (Pearson Education India, 1996).

31. Lynch, M. & Walsh, B. Genetics and Analysis of Quantitative Traits. (Sinauer, 1998).

32. Cai, J., Zhan, J., Arking, D. E. & Bader J. S. Priors, population sizes, and power in genome-wide hypothesis tests. BMC Bioinformatics 24, 170 (2023).

33. Giambartolomei, C. et al. Bayesian Test for Colocalisation between Pairs of Genetic Association Studies Using Summary Statistics. PLOS Genet. 10, e1004383 (2014).

34. Schipper, M. et al. Prioritizing effector genes at trait-associated loci using multimodal evidence. Nat. Genet. 57, 323–333 (2025).

35. Grotzinger, A. D., Fuente J. de la, Privé, F., Nivard, M. G. & Tucker-Drob, E. M. Pervasive Downward Bias in Estimates of Liability-Scale Heritability in Genome-wide Association Study Meta-analysis: A Simple Solution. Biol. Psychiatry 93, 29–36 (2023).

36. Watanabe, K., Taskesen, E., Van Bochoven, A. & Posthuma, D. Functional mapping and annotation of genetic associations with FUMA. Nat. Commun. 8, (2017).

37. Shadrin, A. A. et al. Distinct patterns of genetic overlap among multimorbidities revealed with trivariate MiXeR. Genome Med. 17, 106 (2025).

38. Auton, A. et al. A global reference for human genetic variation. Nature 526, 68–74 (2015).

39. Holland, D. et al. Beyond SNP heritability: Polygenicity and discoverability of phenotypes estimated with a univariate Gaussian mixture model. PLoS Genet. 16, (2020).

40. Altshuler, D. M. et al. Integrating common and rare genetic variation in diverse human populations. Nature 467, 52–58 (2010).

41. Werme, J., van der Sluis, S., Posthuma, D. & de Leeuw, C. A. An integrated framework for local genetic correlation analysis. Nat. Genet. 54, 274–282 (2022).

42. Uffelmann, E., Leeuw, C. de & Posthuma, D. Local Genetic Sex Differences in Quantitative Traits. 2023.05.04.539410 Preprint at 10.1101/2023.05.04.539410 (2023).

43. Werme, J. et al. Local genetic correlation analysis links depression with molecular and brain imaging endophenotypes. medRxiv https://doi.org/10.1101/2023.03.01.23286613 (2023) xdoi:10.1101/2023.03.01.23286613.

44. Tissink, E. et al. The Genetic Architectures of Functional and Structural Connectivity Properties within Cerebral Resting-State Networks. eNeuro 10, (2023).

45. Pullin, J. M. & Wallace, C. Variant-specific priors in colocalisation analysis. 2024.08.21.608957 Preprint at 10.1101/2024.08.21.608957 (2024).

46. Wallace, C. A more accurate method for colocalisation analysis allowing for multiple causal variants. PLOS Genet. 17, e1009440 (2021).

47. A global reference for human genetic variation. Nature 526, 68–74 (2015).

48. Carbon, S. et al. The Gene Ontology resource: Enriching a GOld mine. Nucleic Acids Res. 49, D325– D334 (2021).

49. Knox, C. et al. DrugBank 6.0: the DrugBank Knowledgebase for 2024. Nucleic Acids Res. 52, D1265–D1275 (2024).

50. Frei, O. et al. Bivariate causal mixture model quantifies polygenic overlap between complex traits beyond genetic correlation. Nat. Commun. 10, (2019).

51. Bulik-Sullivan, B. et al. An atlas of genetic correlations across human diseases and traits. Nat. Genet. 47, 1236–1241 (2015).

52. Lee, P. H. et al. Genomic Relationships, Novel Loci, and Pleiotropic Mechanisms across Eight Psychiatric Disorders. Cell 179, 1469–1482.e11 (2019).

53. Prather, A. A., Vogelzangs, N. & Penninx, B. W. J. H. Sleep duration, insomnia, and markers of systemic inflammation: Results from the Netherlands Study of Depression and Anxiety (NESDA). J. Psychiatr. Res. 60, 95–102 (2015).

54. Han, V. X., Patel, S., Jones, H. F. & Dale, R. C. Maternal immune activation and neuroinflammation in human neurodevelopmental disorders. Nat. Rev. Neurol. 17, 564–579 (2021).

55. Zhang, Y. et al. The emerging role of furin in neurodegenerative and neuropsychiatric diseases. Transl. Neurodegener. 11, 39 (2022).

56. Sytnyk, V., Leshchyns’ka, I. & Schachner, M. Neural Cell Adhesion Molecules of the Immunoglobulin Superfamily Regulate Synapse Formation, Maintenance, and Function. Trends Neurosci. 40, 295–308 (2017).

57. Shannon, P. et al. Cytoscape: A software Environment for integrated models of biomolecular interaction networks. Genome Res. 13, 2498–2504 (2003).

58. Merico, D., Isserlin, R., Stueker, O., Emili, A. & Bader, G. D. Enrichment map: A network-based method for gene-set enrichment visualization and interpretation. PLoS ONE 5, (2010).

59. Reimand, J. et al. Pathway enrichment analysis and visualization of omics data using g:Profiler, GSEA, Cytoscape and EnrichmentMap. Nat. Protoc. 14, 482–517 (2019).

60. Buniello, A. et al. Open Targets Platform: facilitating therapeutic hypotheses building in drug discovery. Nucleic Acids Res. 53, D1467–D1475 (2025).

61. Watanabe, K. et al. Genome-wide meta-analysis of insomnia prioritizes genes associated with metabolic and psychiatric pathways. Nat. Genet. 54, 1125–1132 (2022).

62. Mackay, T. F. C. & Anholt R. R. H. Pleiotropy, epistasis and the genetic architecture of quantitative traits. Nat. Rev. Genet. 1–19 (2024) doi:10.1038/s41576-024-00711-3.

63. Cutler, A. J., Mattingly, G. W. & Maletic, V. Understanding the mechanism of action and clinical effects of neuroactive steroids and GABAergic compounds in major depressive disorder. Transl. Psychiatry 13, 228 (2023).

64. Krystal, J. H., Kavalali, E. T. & Monteggia, L. M. Ketamine and rapid antidepressant action: new treatments and novel synaptic signaling mechanisms. Neuropsychopharmacology 49, 41–50 (2024).

65. Lucassen, P. J. et al. Regulation of adult neurogenesis by stress, sleep disruption, exercise and inflammation: Implications for depression and antidepressant action. Eur. Neuropsychopharmacol. 20, 1–17 (2010).

66. Duman, R. S., Sanacora, G. & Krystal, J. H. Altered Connectivity in Depression: GABA and Glutamate Neurotransmitter Deficits and Reversal by Novel Treatments. Neuron 102, 75–90 (2019).

67. Duman, R. S., Aghajanian, G. K., Sanacora, G. & Krystal, J. H. Synaptic plasticity and depression: new insights from stress and rapid-acting antidepressants. Nat. Med. 22, 238–249 (2016).

68. Bisaz, R., Conboy, L. & Sandi, C. Learning under stress: A role for the neural cell adhesion molecule NCAM. Neurobiol. Learn. Mem. 91, 333–342 (2009).

69. Manitt, C. et al. dcc orchestrates the development of the prefrontal cortex during adolescence and is altered in psychiatric patients. Transl. Psychiatry 3, e338–e338 (2013).

70. Kleijer, K. T. E., Zuko, A., Shimoda, Y., Watanabe, K. & Burbach, J. P. H. Contactin-5 expression during development and wiring of the thalamocortical system. Neuroscience 310, 106–113 (2015).

71. Kim, J. et al. MDGA1 negatively regulates amyloid precursor protein–mediated synapse inhibition in the hippocampus. Proc. Natl. Acad. Sci. 119, e2115326119 (2022).

72. Katzman, A. & Alberini, C. M. NLGN1 and NLGN2 in the prefrontal cortex: their role in memory consolidation and strengthening. Curr. Opin. Neurobiol. 48, 122–130 (2018).

73. Babaev, O. et al. IgSF9b regulates anxiety behaviors through effects on centromedial amygdala inhibitory synapses. Nat. Commun. 9, 5400 (2018).

74. Hruska, M. & Dalva, M. B. Ephrin regulation of synapse formation, function and plasticity. Mol. Cell. Neurosci. 50, 35–44 (2012).

75. Lie, E., Li, Y., Kim, R. & Kim, E. SALM/Lrfn Family Synaptic Adhesion Molecules. Front. Mol. Neurosci. 11, (2018).

76. Kim, S., Shin, J. J., Kang, M., Yi, Y. & Kim, E. Synaptic and Non-Synaptic Functions of PTPRD: A Receptor Tyrosine Phosphatase at the Crossroads of Neural Circuitry and Metabolism. J. Neurochem. 169, e70292 (2025).

77. Sha, Z., Xu, J., Li, N. & Li, O. Regulatory Molecules of Synaptic Plasticity in Anxiety Disorder. Int. J. Gen. Med. 16, 2877–2886 (2023).

78. Zhang, M.-Q., Li, R.Wang, Y.-Q. & Huang, Z.-L. Neural Plasticity Is Involved in Physiological Sleep, Depressive Sleep Disturbances, and Antidepressant Treatments. Neural Plast. 2017, 5870735 (2017).

79. Smoller, J. W., Lunetta, K. L. & Robins, J. Implications of comorbidity and ascertainment bias for identifying disease genes. Am. J. Med. Genet. 96, 817–822 (2000).

80. Kendler, K. S., Chatzinakos, C. & Bacanu, S.-A. The impact on estimations of genetic correlations by the use of super-normal, unscreened, and family-history screened controls in genome wide case– control studies. Genet. Epidemiol. 44, 283–289 (2020).

81. Lee, P. H., Feng, Y.-C. A. & Smoller, J. W. Pleiotropy and Cross-Disorder Genetics Among Psychiatric Disorders. Biol. Psychiatry 89, 20–31 (2021).

