## Supplementary_notes for "Shared predicted effector genes of insomnia, anxiety and depression implicate synaptic processes as transdiagnostic drug targets"

### 1. Phenotype definitions in GWAS analysed in this study

The phenotypes used in this study have been constructed using available phenotypic data, and can vary from clinical diagnoses to self-reported diagnoses to approximations of these disorders based on available data. Here we provide a brief overview of the data used to construct the phenotypes used for the GWAS in the studies we analyse in this work.

*Insomnia:* The insomnia GWAS is comprised of two main cohorts, the UK biobank (UKB) and 23andMe. The UKB phenotype is constructed based on self-reported problems falling or staying asleep, where "often" is scored as case, and "sometimes" or "never" is scored as control. The 23andMe phenotype is constructed of 7 distinct datapoints, comprising of self-reported sleep problems and clinical diagnoses. The first large-scale GWAS of insomnia extensively validated this phenotype definition[1] against the gold-standard clinical diagnoses of Insomnia Disorder in the Netherlands Sleep Registry. In short, when assessed via the Duke Structured Interview for Sleep Disorders (DSM-5 and ICSD-3 criteria) and validated Insomnia Severity Index (ISI) and the Pittsburgh Sleep Quality Index (PSQI) thresholds, the single-item question on trouble falling or staying asleep was able to discriminate insomnia disorder cases from controls with outstanding sensitivity, specificity and accuracy (all >.89). Its accuracy across all 19 ICD-10 categories (including mental and behavioral disorders) ranged significantly lower (.57-.63). For more information, see Supplementary Text of Hammerschlag et al. (2017).

*Depression:* Depression cases were defined through clinical diagnoses in electronic health records in Danish (iPSYCH) and Finnish (FinnGen) health registries, and based on diagnoses in electronic health records in the Million Veterans Program (MVP), and self-reported clinical diagnoses (UKB).

*Anxiety:* Anxiety phenotype definitions ranged from narrow diagnostic anxiety disorders (Psychiatric Genetics Consortium [PGC]) to broad anxiety-related disorders (iPSYCH, FinnGen, UKB, All of Us) and symptom-level anxiety severity (MVP). Cases can be defined through a broad range of data, from clinical diagnoses from health registries and electronic health records and self-reported anxiety.

**2. Step-by-step FLAMES methodology**

FLAMES (Fine-mapped Locus Assessment Model of Effector geneS)[2] consists of five steps. First, each locus was fine-mapped to obtain 95% credible sets. We ran PolyFun (POLYgenic FUNctionally-informed fine-mapping)[3] on each locus with default settings except for keeping SNPs in the HLA region. We extracted the estimated prior causal probabilities for the SNPs, which were then used by SuSiE (Sum of Single Effects)[4] to perform non-functionally informed fine-mapping assuming one causal variant per locus. Second, we ran MAGMA[5] as implemented in the CoMorMent Containers v1.8.1[6] to obtain gene-level *z*-scores. We then ran MAGMA tissue type analysis using these *z*-scores on the preformatted GTEx[7] tissue expression file provided by the FLAMES authors. Third, Polygenic Priority Score (PoPS; v0.2)[8] was used to compute convergence-based gene prioritization scores. The FLAMES authors provided PoPS features including an extensive set of public bulk and single-cell expression datasets and predicted protein-protein interactions (https://zenodo.org/records/12635505). We intentionally excluded biological pathways in the PoPs features, aiming to prevent circularity in downstream pathway analyses on FLAMES prioritized genes[2]. Fourth, we annotated credible sets with FLAMES annotate, which creates gene-level evidence scores for genes within 750kb of the centroids of credible sets from SNP-to-gene annotations, MAGMA Z-scores and PoPS scores. We used the FLAMES predictive model to predict the effector gene of each credible set from the generated annotations.

**3. Genome-wide (partial) genetic correlations**

We computed pairwise global genetic correlations ($r_g$) between INS, ANX and DEP summary statistics (EUR) using LD Score Regression (LDSC)[9]. 1000 Genomes EUR[10] served as reference panel. LDSC analysis is restricted to HapMap3 SNPs[11], ensuring a comparable set of SNPs available across traits with global genome coverage. The results (Supplementary Figure 1; $r_{g_{\text{INS-DEP}}}$ = .63 [SE = .01], $r_{g_{\text{INS-ANX}}}$ = 0.63 [SE = .02]; $r_{g_{\text{DEP-ANX}}}$ = .91 [SE = .01]) were in line with previous studies[12,13].

We additionally applied partial LDSC[14] with the same input and settings, which is an R-package extending standard $r_g$ from LDSC (https://github.com/GEMINI-multimorbidity/partialLDSC). We used this tool to condition the $r_g$ between DEP and ANX on INS, between INS and DEP on ANX, and between INS and DEP on ANX. We found that accounting for the SNP effects of INS significantly reduces the genetic correlation between DEP and ANX to $r_{g_{\text{DEP-ANX | INS}}}$ = .87 (SE = .04, $T_{\text{diff}} = 3.27$, $p_{\text{diff}} = 1.07\times10^{-3}$). As expected from the strong global $r_g$ between DEP and ANX, conditioning DEP-INS on ANX and ANX-INS on DEP greatly reduces the found genetic correlation ($r_{g_{\text{INS-ANX | DEP}}}$ = .17 [SE = .08]; $r_{g_{\text{INS-DEP | ANX}}}$ = .16 [SE = .08]).

To assess the influence of phenotype ascertainment and cohort overlap, we repeated the LDSC and partial LDSC analyses using DEP summary statistics based on the narrow definition (excluding the UKB broad depression self-report), INS summary statistics excluding UKB (23andMe only), and ANX summary statistics excluding UKB. Genetic correlations were similar to the primary analysis: $r_{g_{\text{DEP-ANX}}}$ = .97 (SE = .01), $r_{g_{\text{INS-DEP}}}$ = .65 (SE = .02), $r_{g_{\text{INS-ANX}}}$ = .62 (SE = .02). Conditioning on INS changed the DEP-ANX correlation little ($r_{g_{\text{DEP-ANX | INS}}}$ = .95, SE = .03). Conditioning INS-ANX on DEP and INS-DEP on ANX attenuated these correlations to $r_{g_{\text{INS-ANX | DEP}}}$ = −.09 (SE = .14) and $r_{g_{\text{INS-DEP | ANX}}}$ = .29 (SE = .11), consistent with the primary analysis given the near-collinearity of DEP and ANX. These analyses reduce but do not eliminate sample overlap and ascertainment effects, since the remaining cohorts contribute to more than one trait. They also cannot address diagnostic criteria shared between the disorders, so the estimated overlap should be interpreted as shared genetic liability of the co-occurring phenotypes. Disorder-specific designs that exclude comorbid cases may be more informative about distinct biology, as shown for ANX and DEP[15].

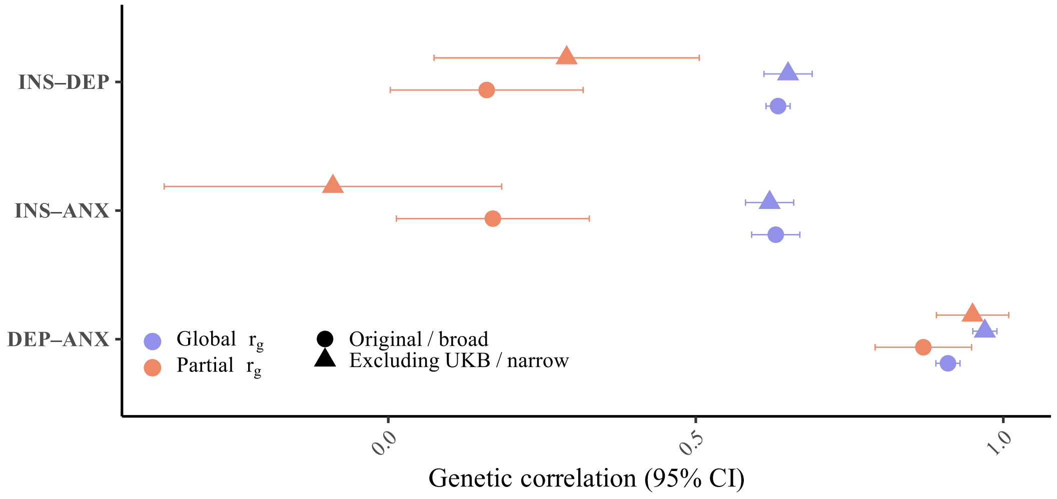


*Supplementary Figure 1.* **Pairwise genetic correlations between INS, DEP and ANX.** Global $r_g$ was based on LDSC, partial $r_g$ between two traits depends on conditioning on the third trait (error bars indicate 95% CI).

### 4. Colocalization

The percentage of significant colocalization was higher (62%) in the GWS loci than in the LAVA loci (42%), which is a significant difference (Fisher's exact test, $p = 1.5\times10^{-3}$). This discrepancy can be explained by (1) LAVA loci generally being larger than GWS loci allowing for different signals residing within the same region, and/or (2) a lack of power. All our LAVA loci were significant in the univariate test of local heritability ($p_{\text{univ}} < 1\times10^{-4}$), yet for these loci, the COLOC posterior probability for at least one trait having no signal for a causal variant in the locus is, on average, 34% compared to 7% for GWS significant loci. Of the remaining posterior probability, on average, 64% of the probability points to colocalization, whereas 36% of the probability points to non-colocalizing signal for both traits. These probabilities are highly similar (65% and 35% respectively) for GWS loci. This might indicate that the COLOC analysis based on LAVA loci is hindered by statistical power, and the colocalization estimate in the LAVA loci might be too conservative.

Across all loci (GWS and LAVA), the average proportion of colocalization probability is 64% vs. 36% non-colocalization probability when signal is detected for both traits. This suggests that the signal observed in about two thirds of the overlapping GWS and LAVA loci is consistent with a shared causal variant.

### 5. Effector genes shared by INS and ANX

Only two genes overlap between INS and ANX without also being mapped to DEP. For statistical reasons we did not test these two genes, as power is extremely limited for such a small set. Nevertheless, their functions align with the significantly enriched gene-sets. In rodents, acute stress upregulates *PDE4B* within a defined GABAergic projection, reducing postsynaptic cyclic adenosine monophosphate (cAMP) signalling and producing anxiety-like behaviour. This effect can be reversed by projection-specific *Pde4b* knockdown in the same circuit[16]. Pharmacological *PDE4D* inhibition also mitigates sleep deprivation-induced memory impairments[17]. *FTO* is known to be involved in the regulation of dopaminergic midbrain circuitry[18].

### 6. Tissue and cell-type enrichments of shared gene-sets

We performed tissue-type and cell-type specificity analysis using FUMA'S GENE2FUNC[19], for three sets of shared predicted effector genes. We find significant enrichments for brain tissue generally (Supplementary Figure 2), with neuronal cell-types being most strongly associated in an enrichment analysis of cell-type markers using the MSigDB c8 dataset (Supplementary Figure 3).

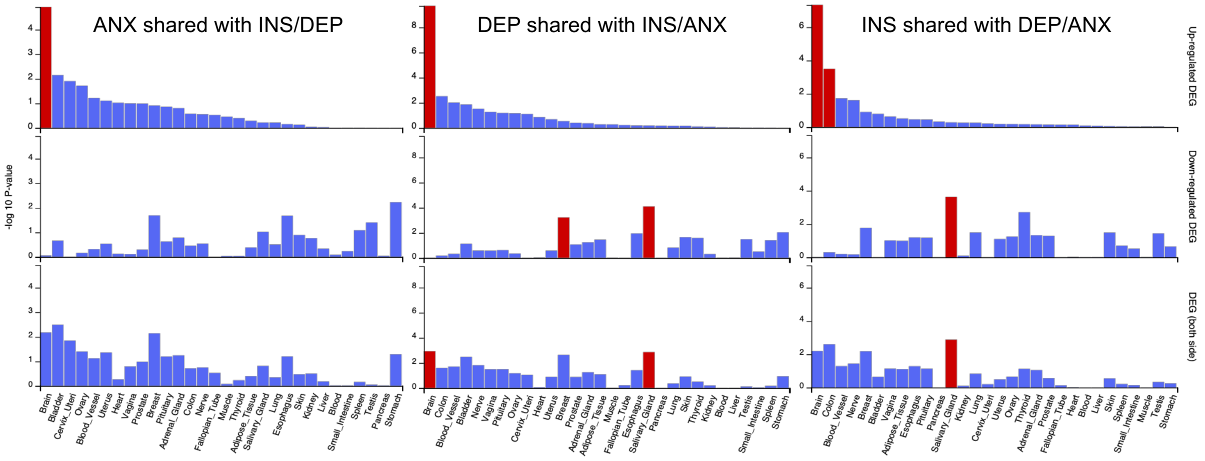


*Supplementary Figure 2.* **Predicted effector genes shared across INS, ANX and DEP show upregulated expression in brain tissue.** Based on Genotype-Tissue Expression (GTEx)[20] data.

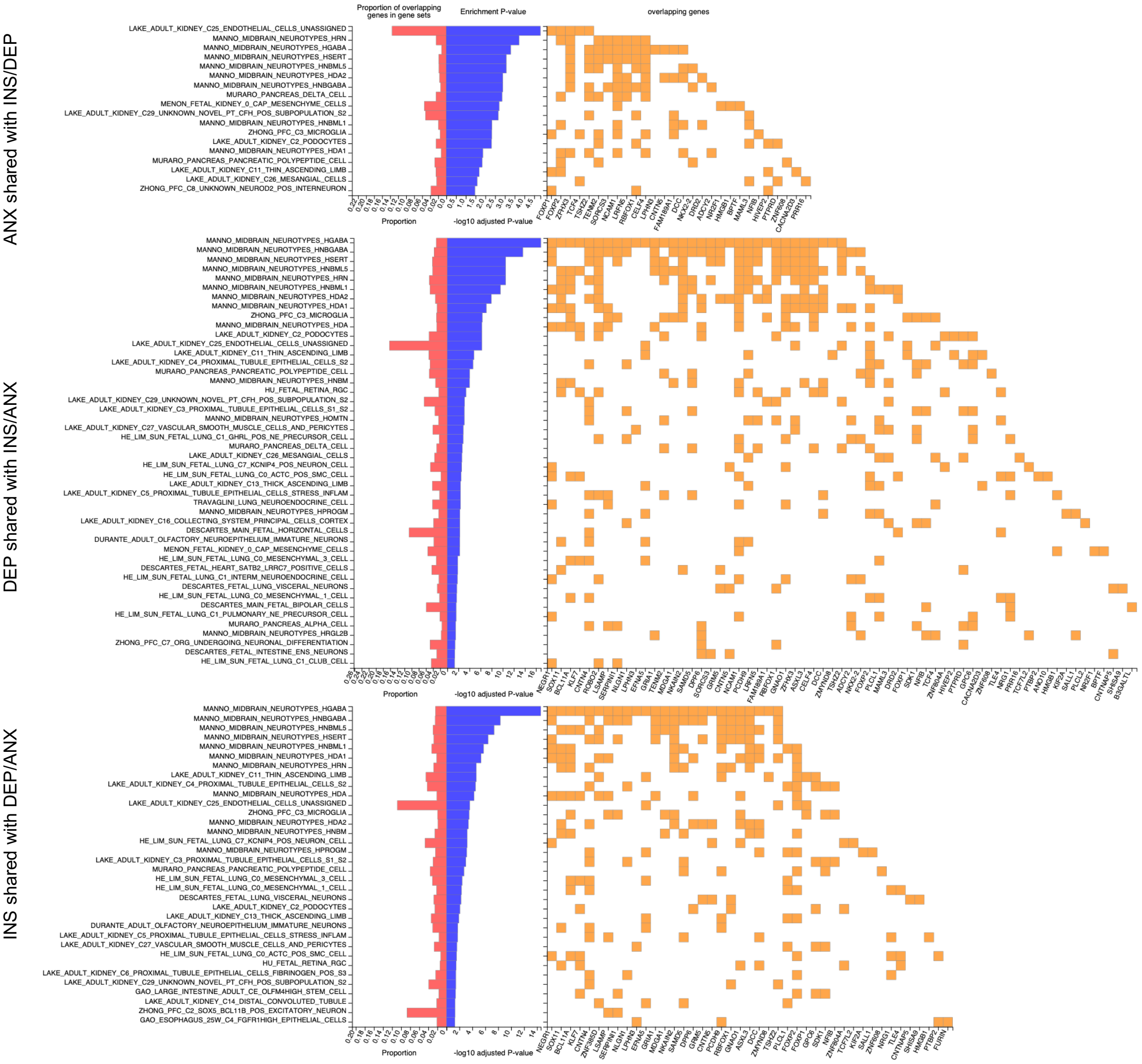


*Supplementary Figure 3.* **Predicted effector genes shared across INS, ANX and DEP are enriched in markers for neuronal cell-types.** Based on MSigDB c8 dataset[21].

**7. Divergence between FLAMES and COLOC**

The prioritization of different predicted effector genes in the presence of evidence for a shared causal variant could be a result of difference in overall fine-mapped credible sets despite significant colocalization, which could be a consequence of the single variant assumption used. Alternatively, these differences could be due to different weighting of eQTL evidence based on tissue relevance weighting step in FLAMES based on genome-wide MAGMA scores. The differences could also be due to different PoPS scores for genes in the locus between these traits, which could capture tissue/cell-type relevance of the GWAS signal based on the large quantity of tissue-specific co-expression networks included in the features.

### References


1. Hammerschlag, A. R. *et al.* Genome-wide association analysis of insomnia complaints identifies risk genes and genetic overlap with psychiatric and metabolic traits. *Nat. Genet.* **49**, 1584–1592 (2017).
2. Schipper, M. *et al.* Prioritizing effector genes at trait-associated loci using multimodal evidence. *Nat. Genet.* **57**, 323–333 (2025).
3. Weissbrod, O. *et al.* Functionally-informed fine-mapping and polygenic localization of complex trait heritability. https://doi.org/10.1101/807792 (2020) doi:10.1101/807792.
4. Wang, G., Sarkar, A., Carbonetto, P., Stephens, M. & Matthew Stephens, C. A simple new approach to variable selection in regression, with application to genetic fine-mapping. https://doi.org/10.1101/501114 doi:10.1101/501114.
5. de Leeuw, C. A., Mooij, J. M., Heskes, T. & Posthuma, D. MAGMA: Generalized Gene-Set Analysis of GWAS Data. *PLoS Comput. Biol.* **11**, (2015).
6. Akdeniz, B. C. *et al.* COSGAP: COntainerized Statistical Genetics Analysis Pipelines. *Bioinforma. Adv.* **4**, vbae067 (2024).
7. Lonsdale, J. *et al.* The Genotype-Tissue Expression (GTEx) project. *Nat. Genet.* **45**, 580–585 (2013).
8. Weeks, E. M. *et al.* Leveraging polygenic enrichments of gene features to predict genes underlying complex traits and diseases.
9. Bulik-Sullivan, B. *et al.* An atlas of genetic correlations across human diseases and traits. *Nat. Genet.* **47**, 1236–1241 (2015).
10. Auton, A. *et al.* A global reference for human genetic variation. *Nature* **526**, 68–74 (2015).
11. Altshuler, D. M. *et al.* Integrating common and rare genetic variation in diverse human populations. *Nature* **467**, 52–58 (2010).
12. Romero, C. *et al.* Exploring the genetic overlap between twelve psychiatric disorders. *Nat. Genet.* **54**, 1795–1802 (2022).
13. Grotzinger, A. D. *et al.* Genetic architecture of 11 major psychiatric disorders at biobehavioral, functional genomic and molecular genetic levels of analysis. *Nat. Genet.* **54**, 548–559 (2022).
14. Mounier, N. *et al.* Genetics identifies obesity as a shared risk factor for co-occurring multiple long-term conditions. 2024.07.10.24309772 Preprint at https://doi.org/10.1101/2024.07.10.24309772 (2024).
15. Tesfaye, M. *et al.* Comorbidity Alters the Genetic Relationship Between Anxiety Disorders and Major Depression. *Biol. Psychiatry* **100**, 689–698 (2026).
16. Xiao, Z.-X. *et al.* Pde4b-regulated cAMP signaling pathway in the AUDGABA-S1TrSst circuit underlies acute-stress-induced anxiety-like behavior. *Cell Rep.* **44**, (2025).

17. Zhao, H., Blokland, A., Prickaerts, J., Havekes, R. & Heckman, P. R. A. Treatment with the selective PDE4B inhibitor A-33 or PDE4D inhibitor zatolmilast prevents sleep deprivation-induced deficits in spatial pattern separation. *Behav. Brain Res.* **459**, 114798 (2024).

18. Hess, M. E. *et al.* The fat mass and obesity associated gene (Fto) regulates activity of the dopaminergic midbrain circuitry. *Nat. Neurosci.* **16**, 1042–1048 (2013).

19. Watanabe, K., Taskesen, E., van Bochoven, A. & Posthuma, D. Functional mapping and annotation of genetic associations with FUMA. *Nat. Commun.* **8**, 1826 (2017).

20. THE GTEX CONSORTIUM. The GTEx Consortium atlas of genetic regulatory effects across human tissues. *Science* **369**, 1318–1330 (2020).

21. Liberzon, A. *et al.* Molecular signatures database (MSigDB) 3.0. *Bioinformatics* **27**, 1739–1740 (2011).
